# Women in Health Care Experiencing Occupational Stress and Burnout during COVID-19: A Review

**DOI:** 10.1101/2021.01.08.21249468

**Authors:** Abi Sriharan, Savithiri Ratnapalan, Andrea C. Tricco, Doina Lupea

## Abstract

**Context:** COVID-19 has had an unprecedent impact on physicians, nurses, and other health professionals around the world, and a serious health care burnout crisis is emerging as a result of this pandemic.

**Objectives:** We aim to identify the causes of occupational stress and burnout in women in medicine, nursing, and other health professions during the COVID-19 pandemic and interventions that can support female health professionals deal with this crisis through a rapid review.

**Methods:** We searched MEDLINE, Embase, CINAHL, PsycINFO, and ERIC from December 2019 through September 30, 2020. The review protocol was registered in PROSPERO and is available online. We selected all empirical studies that discussed stress and burnout in women health care workers during the COVID-19 pandemic.

**Results:** The literature search identified 6148 citations. A review of abstracts led to the retrieval of 721 full-text articles for assessment, of which 47 articles were included for review. Our findings show that concerns of safety (65%), staff and resource adequacy (43%), workload and compensation (37%), job roles and security (41%) appeared as common triggers of stress in the literature.

**Conclusions and Relevance:** The current literature primarily focuses on self-focused initiatives such as wellness activities, coping strategies, reliance of family, friends and work colleagues to organizational led initiatives such as access to psychological support and training. Very limited evidence exists about the organizational interventions such as work modification, financial security, and systems improvement.

## Introduction

The health sector is facing an unprecedent burden due to the ongoing COVID-19 pandemic. Health care workers (HCWs) are at the frontline providing essential services, and they are experiencing increased harassment, stigmatization, physical violence, and psychological trauma, including increased rates of burnout, depression, anxiety, substance abuse, and suicide due to COVID-19^1,2,3,4^. Amnesty International has recorded the deaths of over 7000 health workers worldwide due to COVID-19. In the United States alone, over 250,000 health workers have been infected, and nearly 1000 deaths have occurred ^5,6.^ Women in health care experience specific challenges with adapting to COVID-19–related public health measures, in addition to the preexisting systemic challenges related to workplace gender bias, discrimination, sexual harassment, and inequities ^7.^ The pandemic has taken a disproportionate toll on women in the workplace^8^. Women make up 75% of HCWs globally ^9^.

Female physicians are already more likely than male physicians to experience depression, burnout, and suicidal ideation ^10,11^. On average, women performed 2.5 times of unpaid work per day compared to men as parents and primary caregivers to family members^12^.

In this review, we explore factors that may influence stress and burnout in women health professionals and describe how different type of intervention organizations can offer to support women health professionals.

## METHODS

### Overall Objectives

The overall objectives of this review are to (a) explore the triggers of occupational stress, and burnout faced by women in health care during the COVID-19 pandemic and (b) identify interventions that can support their well-being through a systematic review.

## Materials and Methods

We conducted a rapid review in accordance with the WHO Rapid Review Guide ^13^ and reported using the Preferred Reporting Items for Systematic Reviews and Meta-Analyses (PRISMA). The review protocol was registered in PROSPERO and is available online (CRD42020189750).

### Ethical Considerations

This study used secondary data analysis using published research; therefore, it did not require submission to the Research Ethics Committee.

### Theoretical Model

The WHO classified burnout and occupation stress as an occupational phenomenon ^14^. In this context, we used Bolman and Deal’s (2017) four-frame model of leadership to understand the stress and burnout experienced by women health professionals ^15^. The four-frame model provides an approach to describe organizational issues through four perspectives: structural, human resource, symbolic and political. The structural frame focuses on rules, roles, strategy, policies, technology, and work environment. The human resource frame considers individual needs, skills, and relationships. The political frame examines power, conflict, competition, and organizational politics, and the symbolic frame includes culture, meaning, rituals, and stories.

### Research Questions

The following research questions guided the rapid review: What are the triggers of stress and burnout in women in health care? What interventions are effective in preventing occupational stress and burnout?

### Eligibility Criteria

The eligibility criteria are included in Table 1. First, we were only interested in articles published from December 2019 to September 30, 2020 (the last day of the literature search). We chose this timeframe to include research related to experiences during the COVID-19 pandemic. Our study specifically focused on the experiences of women in health care, encompassing a broad array of health professionals including doctors, nurses, pharmacists, midwives, paramedics, physical therapists, technicians, personnel support workers, and community health workers. We only included articles that focused primarily on women in health care or that provided a breakdown of data according to sex/gender. Given the transboundary nature of the COVID-19 pandemic, we included articles published globally. We defined occupational stress as the degree to which one feels overwhelmed and unable to cope as a result of unmanageable work-related pressures, and we defined burnout as the experience of emotional exhaustion, depersonalization, or cynicism, along with feelings of diminished personal efficacy or accomplishment in the context of the work environment^16^. We included primary where data were collected and analyzed using objective quantitative, qualitative, and mixed methods. We excluded editorials and opinion pieces.

**Table 1:**
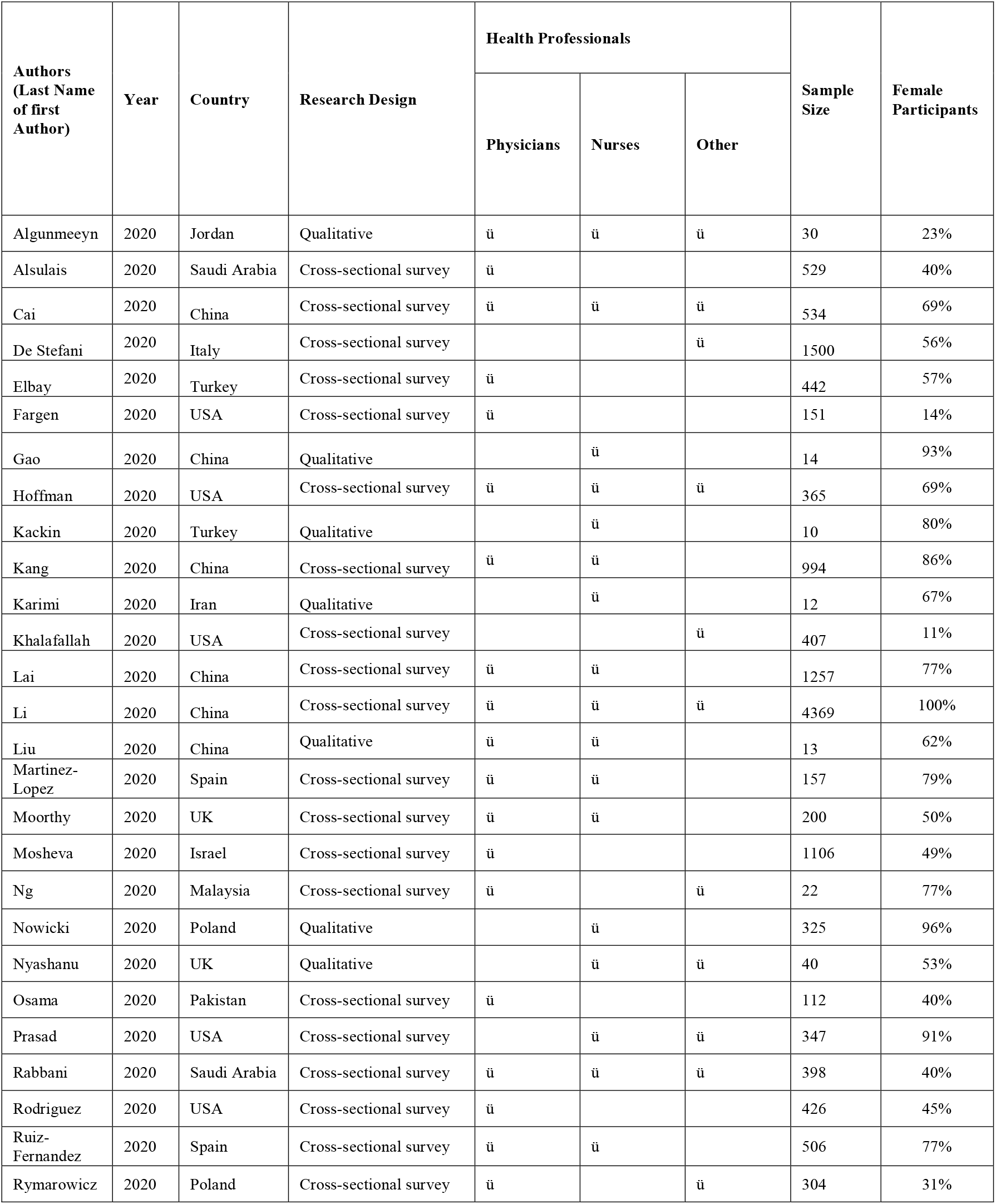
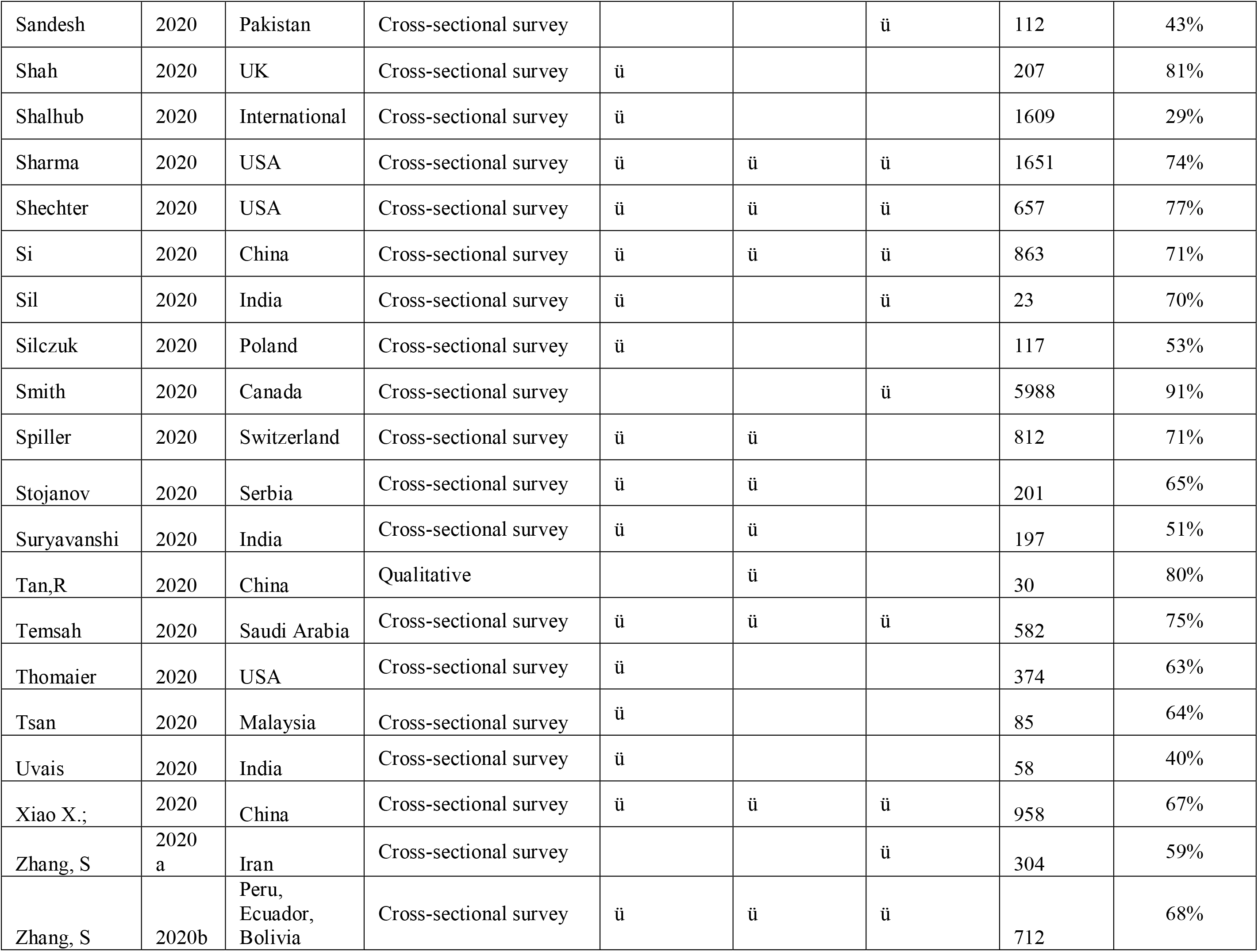
Study Characteristics

### Search Methods and Information Sources

We conducted comprehensive literature search strategies in the following electronic databases: MEDLINE (via Ovid), Embase (via Ovid), CINAHL (via EBSCOhost), PsycINFO (via Ovid), and ERIC (via ProQuest). We developed our search strategies via an academic health sciences librarian with input from the research team. The search was originally built in MEDLINE Ovid and peer-reviewed using the Peer Review of Electronic Search Strategies tool ^17^. We limited our searches to articles published in English no later than September 30, 2020. The final search results were exported into Covidence, review management software, where duplicates were identified and removed.

### Screening Process

To minimize selection bias, we piloted 20 citations against *a priori* inclusion and exclusion criteria. After high agreement was achieved, two reviewers independently screened all citations. Conflicts were resolved by discussion or via a third reviewer. The same process was used for full-text screening of potentially eligible studies.

### Rating of the Quality of Evidence

The strength of data and subsequent recommendations for interventions were graded according to the Quality Rating Scheme for Studies and Other Evidence by two reviewers independently, with discrepancies resolved after joint review and discussion ^18^.

### Data Extraction

We used a predefined data extraction form to extract data from the papers included in the rapid review. To ensure the integrity of the assessment, we piloted the data extraction form on 3 studies. We extracted the following information from the studies: the first author, year of publication, health professionals enrolled in the study, geographic location, study methods, quality of evidence, triggers of stress and burnout, interventions, and outcomes.

### Data Synthesis

Due to heterogeneity of data collected in the included studies, meta-analysis was not appropriate. Instead, we thematically synthesized the data using the thematic analysis process described in Clarke et al. (2012) and grouped the triggers using Bolman and Deal’s (1991) four frame model of leadership^19^.

## Results

### Search Results

The literature search resulted in a total of 6148 records. After 1606 duplicates were removed, 4542 records remained to be screened. We assessed 721 full-text articles and found 47 published studies with 18,668 female health workers met our inclusion criteria. The PRISMA flowchart presents the selection of publications (see Figure 1).

**Figure 1:**
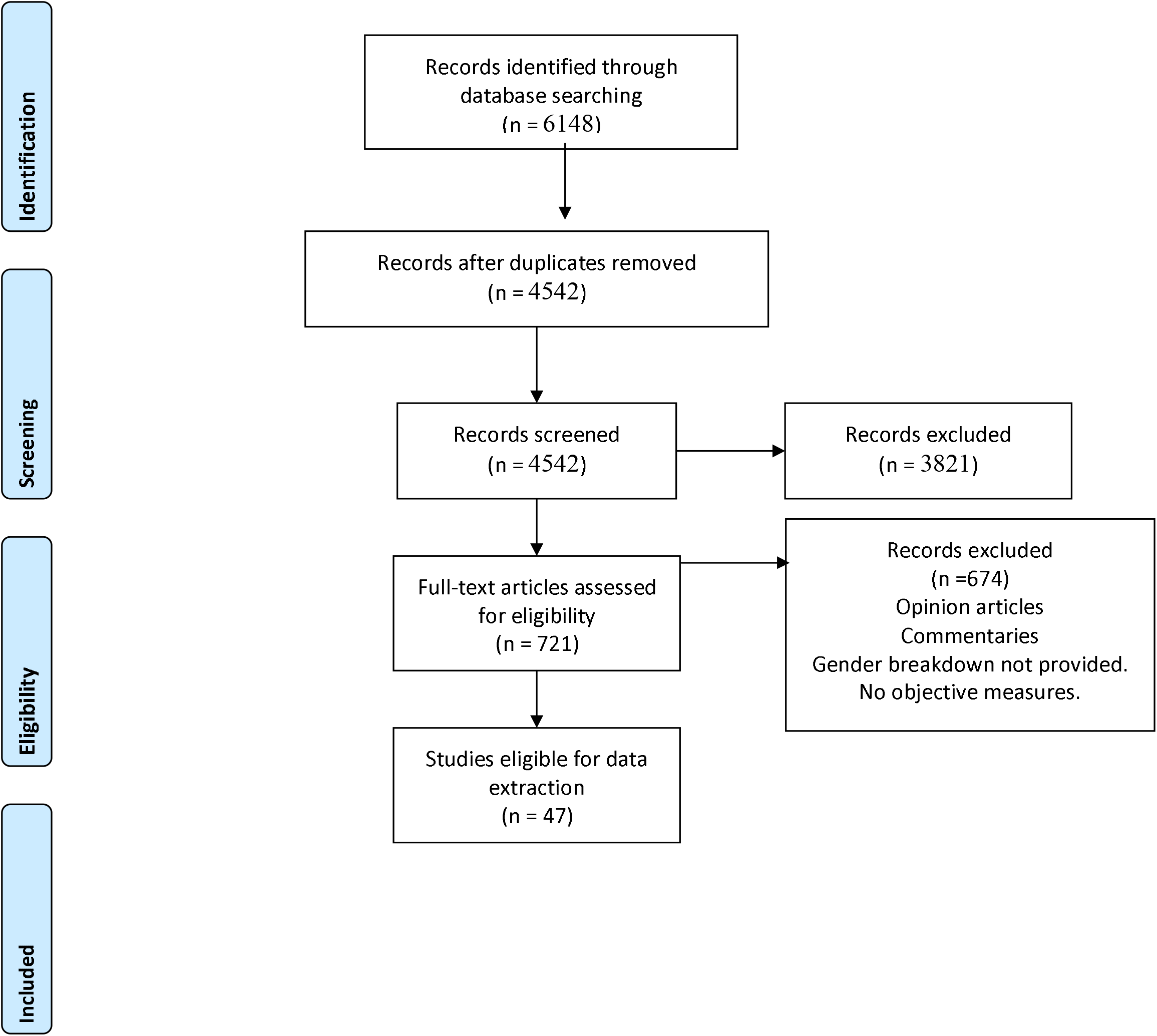
Women in Health Care PRISMA 2009 Flow Diagram.

### Characteristics of Studies

Our search identified 47 eligible studies. Of these, 39 (83%) were cross-sectional studies and 8 (17%) were qualitative studies. Studies came from Asia (34%), Europe (27.6%), Middle East (14.9%), North America (19.1%) and Latin America (2%) (see Table 1). These studies focused on physicians (74%), nurses (57%), and other health professionals (45%; including dentists, personal support workers, pharmacists, and administrative professionals). The study samples often included both male and women health professionals; however, these studies also provided gender-based breakdowns. In all, 62% of the total 29,398 study population focused on female health professionals.

### Triggers of Stress and Burnout Faced by Women in Health Care

Primary forces of stress and burnout in women in health care during COVID-19 were related to structural factors (i.e., organizational resources, work related policies, and roles). Resource adequacy (43%), related to lack of appropriate personal protective equipment (PPE) and staffing shortages, was discussed as a major driver of stress and burnout in the included studies. Stress and burnout intensity differed between health professionals who had indirect patient care and direct clinical care of COVID-19 patients. A total of 43% of the studies reported that caring for COVID-19 patients increased stress and burnout; 38% of the studies reported HCWs faced an increased workload due increased number of COVID-19 patients under their care, and they were not appropriately compensated for the workload.

Regarding the human resource perspective, safety concerns and fears of getting infected with COVID-19 and putting family members at risk (66%) appeared to be the primary causes of stress and burnout. Female gender (34%) and age and family status (19%) also emerged as determinants of risk of stress and burnout. Specifically, being young with no family or being a mother with young children influenced emotional stress and burnout in women. Similarly, less work experience and self-perception about lack of competency to care for COVID-19 patients was associated with increased prevalence of stress and burnout (26%).

In terms of the symbolic frame, concerns about organizational culture (26%), patient care protocols (17%), and societal experiences of health professionals (26%) emerged as common triggers of stress. More specifically, issues related to ambiguous patient care protocols and perceived lack of infection control guidelines influenced stress and burnout. Similarly, the organizational culture, including lack of support and recognition by peers, supervisors, and hospital leadership, were triggers of stress and burnout in women health professionals. From a macro cultural perspective, the societal and media portrayal of health care workers as “heroes” increased moral responsibility and caused increased stress to meet these expectations, yet health professionals faced increased social isolation and stigma as they were considered as contagious by the general population.

From the political perspective, the government-level social distancing protocols increased social isolation (15%). Further, lack of pandemic preparedness (2%), poor public health guidance on screening and treatment (4%), and measures related to infrastructure such as delayed testing and lack of treatment for COVID-19 patients (4%) exacerbated to stress and burnout in women HCWs.

### Interventions That Can Support the Well-Being of Women HCWs during a Pandemic

Only 38.3% studies have examined potential interventions to support women in health care with COVID-19–related stress and burnout. We grouped the interventions on a spectrum ranging from self-focused intervention to systems-focused interventions (see Table 3). 29.7% included studies primarily focused on addressing well-being and resiliency at the individual level. The current literature discussed self-initiated interventions such as regular exercise, wellness activities such as yoga and meditation, faith-based activities, self-help resources, hobbies, psychological services such as therapists, hotlines, and talk therapy as treatment strategies and other adaptive coping mechanisms as useful preventative strategies for women. From a structural perspective, 21.5% of included studies recommended systems-level interventions such as work modifications, ensuring clear communication about policies, providing access to PPE, offering training related to managing COVID-19, instituting measures to support health professionals financially, providing rest areas for sleep and recovery, offering basic physical needs such as food, and including training programs to improve resiliency were considered potential strategies to support women in healthcare during the pandemic.

**Table 2:** Triggers of Stress and Burnout during COVID-19

| Author | Year | TRIGGERS |  |  |  |  |  |  |  |  |  |  |  |  |  |
| --- | --- | --- | --- | --- | --- | --- | --- | --- | --- | --- | --- | --- | --- | --- | --- |
|  |  | STRUCTURAL |  |  | HUMAN RESOURCES |  |  |  | SYMBOLIC |  |  | POLITICAL |  |  |  |
|  |  | Staff and Resource Adequacy | Workload and Compensation | Job Roles and Job Security | Female Gender | Age/Family Status | Safety | Experience | Patient Care Protocols | Societal Expectations | Organization Culture | Public Health Guidance | Infrastructure | Pandemic preparedness | Social Isolation |
| Algunmeeyn | 2020 | ✓ |  |  |  | . | ✓ | ✓ |  |  |  |  |  |  |  |
| AlSulais | 2020 | . |  |  | ✓ | ✓ | ✓ | ✓ |  | ✓ |  |  |  |  |  |
| Cai | 2020 | . | ✓ | ✓ | . | ✓ | ✓ | . |  |  |  |  | ✓ |  |  |
| DeStefani | 2020 | . | . | ✓ | . | . | ✓ | . | ✓ |  |  | ✓ | . |  |  |
| Elbay | 2020 | . | ✓ |  | ✓ | ✓ | . | ✓ |  |  |  |  |  |  | ✓ |
| Fargen | 2020 |  | ✓ | ✓ |  | . | ✓ |  |  |  |  |  |  |  |  |
| Gao | 2020 | ✓ |  | ✓ | . | . | ✓ |  |  |  |  |  |  |  |  |
| Hoffman | 2020 |  | . | . | . | . |  |  |  |  | ✓ |  |  |  |  |
| Kackin | 2020 | ✓ | ✓ | ✓ | . |  | ✓ |  |  | ✓ |  |  |  |  | ✓ |
| Kang | 2020 |  | . | . | . | . | ✓ |  |  |  | . |  |  |  |  |
| Karimi | 2020 | ✓ | ✓ |  |  | . | ✓ |  | ✓ |  |  |  |  |  | ✓ |
| Khalafallah | 2020 |  | ✓ | ✓ | . |  |  |  |  |  |  |  |  |  |  |
| Lai | 2020 | . | . | ✓ | ✓ | . | . | . | . | . |  |  |  |  |  |
| Li | 2020 | . | . | . | ✓ | . | ✓ | ✓ | . | . |  |  |  |  |  |
| Liu | 2020 | ✓ | ✓ | ✓ | . | . | ✓ | ✓ | . | . | ✓ | . |  |  |  |
| Martinez-Lopez | 2020 | ✓ |  |  |  | . | ✓ |  |  |  |  |  |  |  |  |
| Matthewson | 2020 |  |  | ✓ |  |  |  |  |  |  |  |  |  |  |  |
| Moorthy | 2020 | ✓ |  | ✓ | . |  |  |  |  |  |  |  |  |  | ✓ |
| Mosheva | 2020 |  |  |  |  | . | ✓ |  |  |  |  |  |  |  |  |
| Hau Ng | 2020 |  | ✓ |  |  | . | ✓ |  |  | ✓ | ✓ |  |  |  |  |
| Nowicki | 2020 |  |  |  |  | ✓ |  | ✓ |  |  |  |  |  |  |  |
| Nyashanu | 2020 | ✓ |  |  | ✓ |  | ✓ |  | ✓ |  |  | ✓ | ✓ | ✓ |  |
| Osama | 2020 |  |  |  |  |  | ✓ |  |  |  |  |  |  |  |  |
| Prasad | 2020 |  |  | ✓ | . | ✓ |  |  |  | ✓ |  |  |  |  |  |
| Rabbani | 2020 | ✓ |  |  |  |  | ✓ |  |  |  |  |  |  |  |  |
| Rodriguez | 2020 | ✓ |  | ✓ | ✓ |  |  | ✓ |  |  |  |  |  |  |  |
| Rymarowicz | 2020 |  |  |  |  | . | ✓ |  |  |  |  |  |  |  |  |
| Sandesh | 2020 | ✓ | ✓ | ✓ | ✓ | . | ✓ |  |  | ✓ |  |  |  |  |  |
| Shah | 2020 | ✓ | ✓ | ✓ | ✓ |  | ✓ | ✓ | ✓ | ✓ |  |  |  |  |  |
| Shalhub | 2020 | ✓ | ✓ |  | ✓ |  | ✓ |  |  | ✓ |  |  |  |  |  |
| Sharma | 2020 | ✓ | ✓ | ✓ | ✓ |  | ✓ |  | ✓ | ✓ |  |  |  |  |  |
| Shechter | 2020 | ✓ | ✓ |  |  | . | ✓ |  | ✓ |  |  |  |  |  | ✓ |
| Si | 2020 |  |  | ✓ | . | . | ✓ |  |  |  |  |  |  |  |  |
| Sil | 2020 |  |  |  | ✓ |  |  |  |  |  |  |  |  |  |  |
| Silczuk | 2020 |  |  |  | ✓ |  |  |  |  |  |  |  |  |  |  |
| Smith | 2020 | ✓ |  |  | ✓ |  |  |  | ✓ |  |  |  |  |  |  |
| Spiller | 2020 |  | ✓ |  |  |  |  |  |  |  |  |  |  |  |  |
| Stojanov | 2020 |  |  |  | ✓ | ✓ | ✓ |  |  | ✓ |  |  |  |  |  |
| Suryavanshi | 2020 |  | ✓ | ✓ | . | . | ✓ | ✓ | ✓ | ✓ | ✓ |  |  |  | ✓ |
| Tan, R | 2020 | ✓ | ✓ | ✓ |  |  | ✓ | ✓ |  |  |  |  |  |  |  |
| Temsah | 2020 | ✓ |  |  |  | . | ✓ | ✓ |  |  | ✓ |  |  |  |  |
| Thomaier | 2020 | ✓ |  |  | ✓ | . | ✓ | ✓ |  | ✓ |  |  |  |  |  |
| Tsan | 2020 |  | ✓ |  |  |  |  |  |  |  |  |  |  |  |  |
| Uvais | 2020 |  |  |  | ✓ |  |  |  |  | ✓ |  |  |  |  | ✓ |
| Xiao | 2020 | ✓ |  | ✓ |  | ✓ | ✓ |  |  |  |  |  |  |  |  |
| Zhang, S | 2020<br>a |  |  |  |  | ✓ | . |  |  |  |  |  |  |  |  |
| Zhang, S | 2020<br>b |  | ✓ | ✓ |  | ✓ | ✓ |  |  |  | ✓ |  |  |  |  |

**Table 3:** Interventions to Support Stress and Burnout

| Intervention Spectrum | Intervention Type | Example | Evidence Source | Quality of Evidence Strength |
| --- | --- | --- | --- | --- |
| Self-focused | Self-Coping | Normalization | 26 | 4 |
|  |  | Techniques | 50 |  |
|  | Recovery and Resiliency | Yoga, Meditation | 32 | 4 |
|  |  | Relaxation Techniques | 46 |  |
|  |  | Proper Nutrition | 49 |  |
|  |  | Time off | 56 |  |
|  |  | Rest |  |  |
|  | Physical Activities | Sports | 26 | 4 |
|  |  | Exercise | 49 |  |
|  | Hobbies | Sports, Cooking, | 26 | 4 |
|  |  | Movies, | 32 |  |
|  |  | Music | 56 |  |
|  |  | Reading |  |  |
|  | Faith based Activities | Religion | 47 | 4 |
|  |  |  | 49 |  |
|  | Social Networks | Family | 20 | 4 |
|  |  | Friends | 32 |  |
|  |  | Work Colleagues | 37 |  |
|  |  | Virtual Networks | 46 |  |
|  |  |  | 47 |  |
|  |  |  | 50 |  |
|  | Psychological Support | Psychologists | 20 | 4 |
|  |  | Psychiatrist | 24 |  |
|  |  | Group Counselling | 26 |  |
|  |  | Talk therapy | 27 |  |
|  |  |  | 42 |  |
|  |  |  | 46 |  |
|  |  |  | 49 |  |
|  |  |  | 55 |  |
|  |  |  | 56 |  |
|  |  |  | 57 |  |
| Systems-focused | Training | PPE use | 20 | 4 |
|  |  | SARS COV2 virus | 24 |  |
|  |  | Patient Care Protocols | 44 |  |
|  |  | Resiliency | 53 |  |
|  |  |  | 56 |  |
|  | Communication | Transparent | 24 | 4 |
|  |  | communication between | 42 |  |
|  |  | management and | 47 |  |
|  |  | frontline | 64 |  |
|  | Workplace Resources | Access to proper PPE | 20 | 4 |
|  |  | Work coverage | 42 |  |
|  |  | Isolation units | 47 |  |
|  |  | Places for rest and sleep | 53 |  |
|  |  | Childcare | 56 |  |
|  |  |  | 64 |  |
|  | Workplace<br>Incentives | Flexible work policies | 20 | 4 |
|  |  | Compensation | 24 |  |
|  |  |  | 25 |  |
|  |  |  | 26 |  |
|  |  |  | 42 |  |
|  |  |  | 56 |  |
|  | Process Improvement | Rapid testing for patients | 42 | 4 |
|  |  | Improved infection | 53 |  |
|  |  | control protocols |  |  |

However, these studies did not provide evidence on the effectiveness and utility of these interventions in helping women in health care. There was, however, emerging evidence on the use of maladaptive coping mechanisms such as avoidant coping and substance use ^26,47,52^.

## Discussion

In this rapid review, we examined the triggering factors of occupational stress and burnout in women in health care in the context of the COVID-19 pandemic and potential interventions to mitigate these factors. We provided an overview of the evidence and identification of potential variables that influence the mental health well-being of women in healthcare. The current research literature primarily focuses on prevalence of stress, burnout, depression, and anxiety using a cross-sectional approach to show the presence of these elements at a particular point in time. Further, it looks at burnout as an individual issue that can be mitigated by self-help solutions such as coping, yoga, mindfulness, and practicing resilience. However, very weak evidence exists on the effectiveness of these interventions on women in health care (see figure 2).

**Figure 2:**
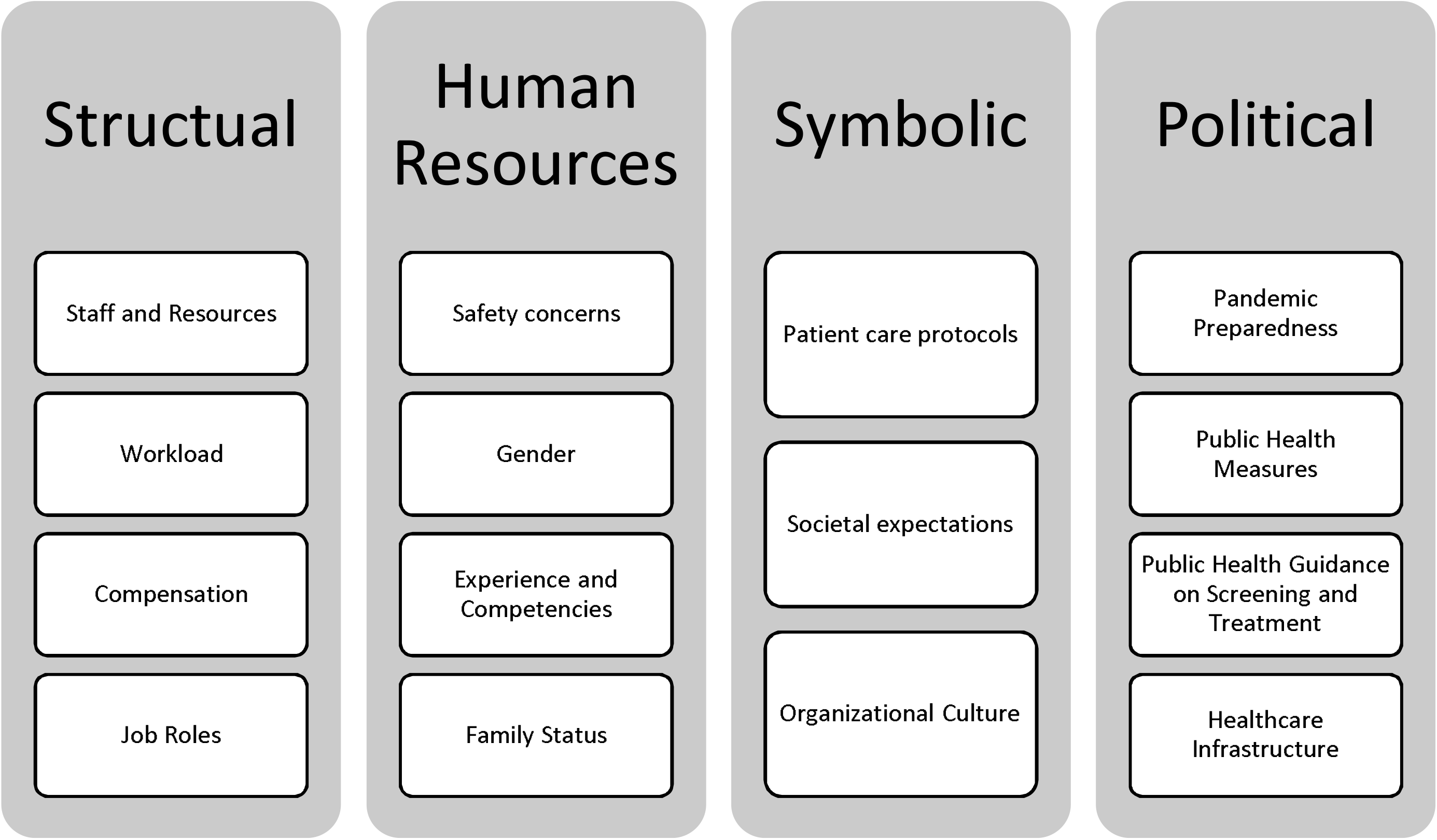
Triggers of Stress and Burnout during COVID-19.

In health care, there is limited understanding about burnout as an occupational phenomenon ^65^. First, there is a gap in the literature regarding how organizations can shape the structures, cultures, and processes to address the elements that triggers of stress and burnout. Similarly, there is a limited understanding of how race, culture, leadership, and profession impact occupational stress and burnout during COVID-19. For example, 1 in 3 nurses who have died of COVID-19 in the United States are from the Filipino community^66^. Similarly, there is a lack of understanding of burnout by occupation type. Physician burnout has received a lot of attention over the past decade, but very limited evidence exists regarding the burnout experienced by other health professionals, including support staff such as personal support workers who are at the frontlines of caring for patients in long-term care and nursing homes.

Similarly, there is very little evidence on how political factors such as policies and public health measures influence individual level burnout. For example, the US Families First Coronovirus Response Act, which required employers to provide up to 80 hours of paid sick leave for reasons related to COVID-19, allowed a provision to exclude health care workers (HCWs) from these benefits. A scan of social media discussions of this showed a significant stress and anxiety among HCWs. Future studies should move beyond cross-sectional studies and explore the contexts, factors, organizational and systems variables, and mechanisms that influence stress and burnout variables to better understand the determinants of stress and burnout in women.

Further, there is very limited evidence on the impact of stress and burnout on quality of care, patient safety, employee engagement, and staff attrition and absenteeism during COVID-19. Future studies on stress and burnout among HCWs should look at the short-, medium-, and long-term impact to health care systems. Specifically, research is needed to understand how COVID-19 will affect women health professional’s decisions about work.

There are several strengths to the current rapid review. To our knowledge, this is the first review that attempted to look at stress and burnout experienced by women in health care as an occupation phenomenon and that explored common triggers of stress and burnout during the COVID-19 pandemic. Our rapid review was guided by the Boleman and Deal’s four-frames theoretical organizational theoretical framework to understand the contextual factors through the lens of structural, human resources, politics, and symbolism. Our methodology was guided by the WHO guidelines on Rapid Reviews and reported using the PRISMA guidelines. The studies included in the review represent a global perspective of the issues. We highlighted the important gap in current understanding related to occupational stress and burnout in Women in Health Care.

The current literature on stress and burnout related to COVID-19 includes both male and female health professionals. Although the studies included in this review provided gender breakdowns in the sample framework and discussed gender related factors, it lacked gender-based subgroup analysis of what interventions are specifically effective for women in health care. Further, our data analysis was limited by the variability of measurement instruments and lack of reporting on structural, political and cultural context of stress and burnout.

There is a significant data gap on the impact of COVID-19 on women in health care. We recommend that national health professional organizations develop comprehensive data gathering and monitoring strategies to improve the science of health professional burnout research.

## Conclusion

Organizational leaders and research scholars should consider occupational stress and burnout as an organizational phenomenon and provide organizational-level support for HCWs. To improve occupational wellness for women in health care, organizations should attempt to engage their health care workforce to listen to their concerns, consider the specific context of the workforce, and design targeted interventions based on their identified needs.

## Data Availability

This review is registered with the Open Science Framework (https://osf.io/y8fdh/?view_only=1d943ec3ddbd4f5c8f6a9290eca2ece7).

## List of Abbreviation

PROSPERO: International database of prospectively registered systematic reviews
HCWs: Health care workers
PRISMA: Preferred Reporting Items for Systematic Reviews and Meta-Analyses
PPE: Personal Protective Equipment

## Declaration

### Ethical Approval and consent to participate

Not required. This study used published data.

### Patients or public

Patients were not involved in the design, or conduct, or reporting, or dissemination plans of our research. The study involved women in health care in the design, conduct, dissemination of our research.

### Consent for publication

Authors give full consent to publish this article. This manuscript has not been published elsewhere.

## Acknowledgement

Authors acknowledge the contribution by Hilary Pang, Ana Patricia Ayala, Dongjoo Lee, Sabine Caleja who helped with article retrieval and screening.

